# Racial and Socioeconomic Fairness of Area-Level Traffic-Related Air Pollution Measure Aggregation

**DOI:** 10.1101/2025.09.26.25336652

**Authors:** Andrew Vancil, Stephen Colegate, Erika Rasnick Manning, Anushka Palipana, Rhonda Szczesniak, Cole Brokamp

## Abstract

**Background:** The Environmental Protection Agency’s Environmental Justice Screen traffic proximity (EJ Screen) and the Department of Transportation’s Average Annual Daily Traffic (AADT) commonly serve as proxies of traffic-related pollution exposure. However, the methods used to aggregate to area-level measures have been untested for bias.

**Methods:** Using a parcel-level measured developed for Hamilton County, Ohio, agreement was determined with both above measures at three geographic levels: census block group, census tract and zip code tabulation area (ZCTA). Fairness was assessed using linear regression.

**Results:** Generally, the values of AADT were in significant agreement with the parcel proximity measure while the EJ Screen was not. Racial and community deprivation bias was widely detected for EJ Screen.

**Discussion:** While the biases detected were not directly against majority black and materially deprived neighborhoods, the biases could manifest in negative downstream effects. These manifestations include suppression of known traffic-related pollution effects in subsequent research.

**Impact Statement:** The Environmental Protection Agency’s Environmental Justice Screen traffic proximity (EJ Screen) and the Department of Transportation’s Average Annual Daily Traffic (AADT) are widely used traffic related pollution proxies however, with common aggregation techniques largely untested for fairness, this research has detected potential biases in the EJ Screen product.

## Background

Traffic related air pollution (TRAP) is detrimental to health and accurately estimating this exposure is key to understanding its effects^1^. TRAP has been linked to a broad spectrum of adverse health outcomes across multiple organ systems and life stages, including harmful effects on respiratory and pulmonary function, cardiovascular health, neurodevelopment and brain function, and others. Epidemiological studies and reviews associate TRAP exposure with increased risks of respiratory diseases (e.g., asthma), cardiovascular disease and related mortality, adverse pregnancy and birth outcomes (e.g. preterm birth), neurocognitive and developmental impairments (e.g., dementia), metabolic disorders (e.g., diabetes), and some cancers.

Epidemiologic studies often rely on proxies of traffic related air pollution because measurements are difficult, even if considered before the collection of health data. Publicly available Average Annual Daily Traffic (AADT) data from United States Department of Transportation ^2,3^ is commonly used as such a proxy. ^2,3^ Area-level averages for traffic air pollution—such as zip codes or census tracts—are widely used due to practical constraints like limited fine-scale monitoring data, computational efficiency, and ease of combining pollution exposure with demographic information for policy decisions^4^. Tools like EPA’s EJScreen exemplify this approach, balancing the trade-off between accuracy at the micro-scale and the necessity of having a consistent, nationally applicable method ^5^. Despite losing some spatial precision, this aggregation enables feasible, timely, and standardized assessment for identifying communities with higher pollution burdens.

However, how AADT estimates are aggregated to area-level estimates and assigned to individuals could introduce exposure misclassification^5^ that is differential with respect to socioeconomic status^6,7^. Studies to date have focused on differential misclassifications of environmental exposures based on geocoding inaccuracies^8–10^, specifically finding that mis-estimation of exposures were associated with area-level socio-economic status.^11^ Other studies have found that different air pollution estimation model resolutions under estimate racial and ethnic exposure disparities^12^ and can often misclassify individual-level exposures^13^. Currently no studies have identified the impacts of exposure aggregation methods on the fairness of exposure assessments.

In this analysis, we seek to evaluate the accuracy and fairness of commonly used area-level AADT estimates, the Environmental Protection Agency’s (EPA) Environmental Justice Screen (EJ Screen)^14^ traffic proximity and an areal measure of AADT density.

## Methods

To evaluate the accuracy and fairness of the areal estimates of traffic proximity, we compared them to the fraction of residential parcels nearby arterial roadways at three different levels of neighborhood: census block group, census tract, and ZIP code tabulation area (ZCTA). We utilized census block group-level EJ Screen estimates pre-calculated as the distance-weighted average annual daily number of vehicles on principal and minor arterial roadways within 500 meters of each block centroid. AADT densities were calculated as the sum of all AADT weighted by the lengths of roadway segments and normalized by the total geographic area.

We utilized tax assessor parcel-level data in Hamilton County, a mixed urban and suburban county in Southwest Ohio to define a measure of the median level of AADT estimated truck traffic on principal and minor arterial roadways roads within 400 m of residential parcels, termed the parcel traffic proximity. In **Figure 1**, we have visualized the three aggregation techniques and what geospatial area is used to generate the traffic estimates. Briefly, **1b** demonstrates how the EJ Screen uses the block group centroid to find included roadways, **1c** depicts the tract-level density calculated of AADT truck traffic, and **1d** demonstrates the generation of the parcel traffic proximity as a function of household size and individual parcel proximity to major roadways. Lastly, traffic estimates were transformed into deciles for comparisons and agreement was defined as “good” if two estimates of traffic differed by no more than one decile. Cohen’s Kappa statistic was used to estimate inter-rater reliability.

**Figure 1:**
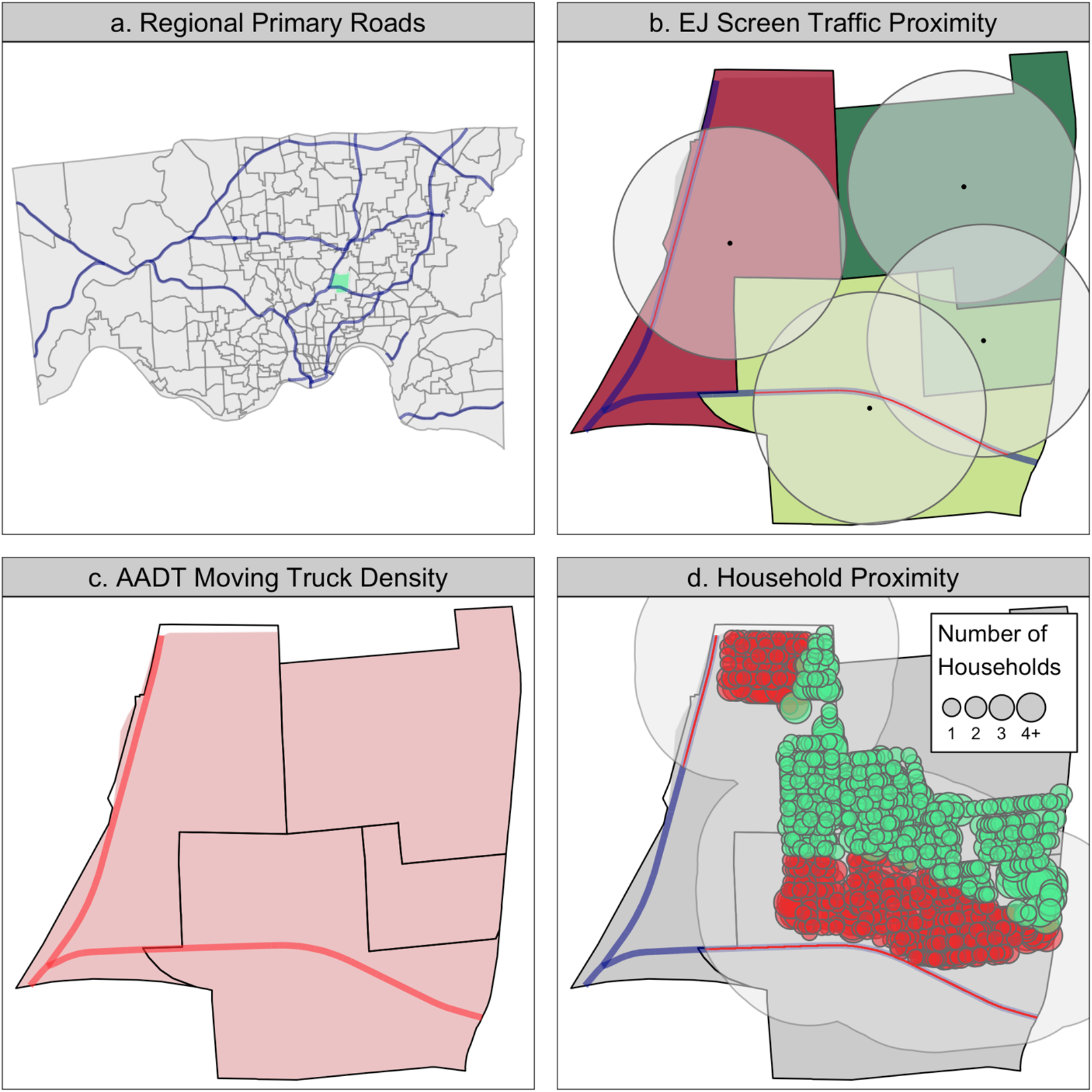
This figure demonstrates the different traffic aggregation techniques. Panel *a* provides an overview of Hamilton County, the primary roads and the example census tract. Panels *b, c*, and *d* visualize the aggregation techniques used in the relevant metrics. Red colored highway sections indicate the portions of highway used in traffic exposure estimate.

Fairness, defined as similarly accurate exposure estimations among populations that have and have not traditionally been marginalized^15^, was operationalized using racial/ethnic composition (fraction non-Hispanic Black) and a census tract-level community material deprivation index^16^. Fairness was visually assessed using neighborhood-level scatter plots and quantified as agreement with the parcel traffic proximity using Kendall’s correlation coefficient on decile-ranked traffic estimates. All analyses were performed using R v4.2.2^17^.

## Results

Our study area was a mixed urban and suburban county in southwest Ohio, consisting of 57 ZCTAs, 222 census tracts and 697 block groups. The county encompasses 2,098 segments of primary roads totaling over 193,394 m with a total AADT of 173,805,990.

Compared to the EJ Screen traffic proximity, the AADT highway truck density measure had higher agreement with the parcel traffic proximity at the block group (72.2% versus 23.7% good agreement) and census tract-level (67.6% versus 37.8%). At the ZCTA-level, both had poor agreement (26.6% versus 33.3%). The AADT measure had higher inter-rater reliability with the parcel traffic proximity compared to the EJ Screen measure (block-group Cohen’s Kappa: 0.17 vs -0.01, tract-level: 0.30 vs 0.13). Both measures had negligible agreement at the ZCTA-level (−0.01 vs 0.05).

At the census-tract level EJ Screen exposure assessment errors were significantly associated with community material deprivation (−0.15, 95% CI: -0.24, -0.05) (**Fig. 2a**) and racial composition (−0.15, 95% CI: -0.24, -0.06) (**Fig. 2b**) while the AADT density errors were not (**Fig. 2c, 2d**). Similarly at the block group level EJ Screen exposure assessment errors were significantly associated with community material deprivation (−0.18, 95% CI: -0.23, -0.13) and racial composition (−0.12, 95% CI: -0.18, -0.07) while the AADT density errors were not. At the ZCTA level, however, the results do suggest estimate bias for the EJ Screen across community material deprivation (−0.31, 95% CI: -0.43, -0.12) and both products across racial composition (EJ Screen: -0.42, 95% CI: -0.56, -0.24; AADT: -0.34, 95% CI: -0.47, -0.16).

**Figure 2:**
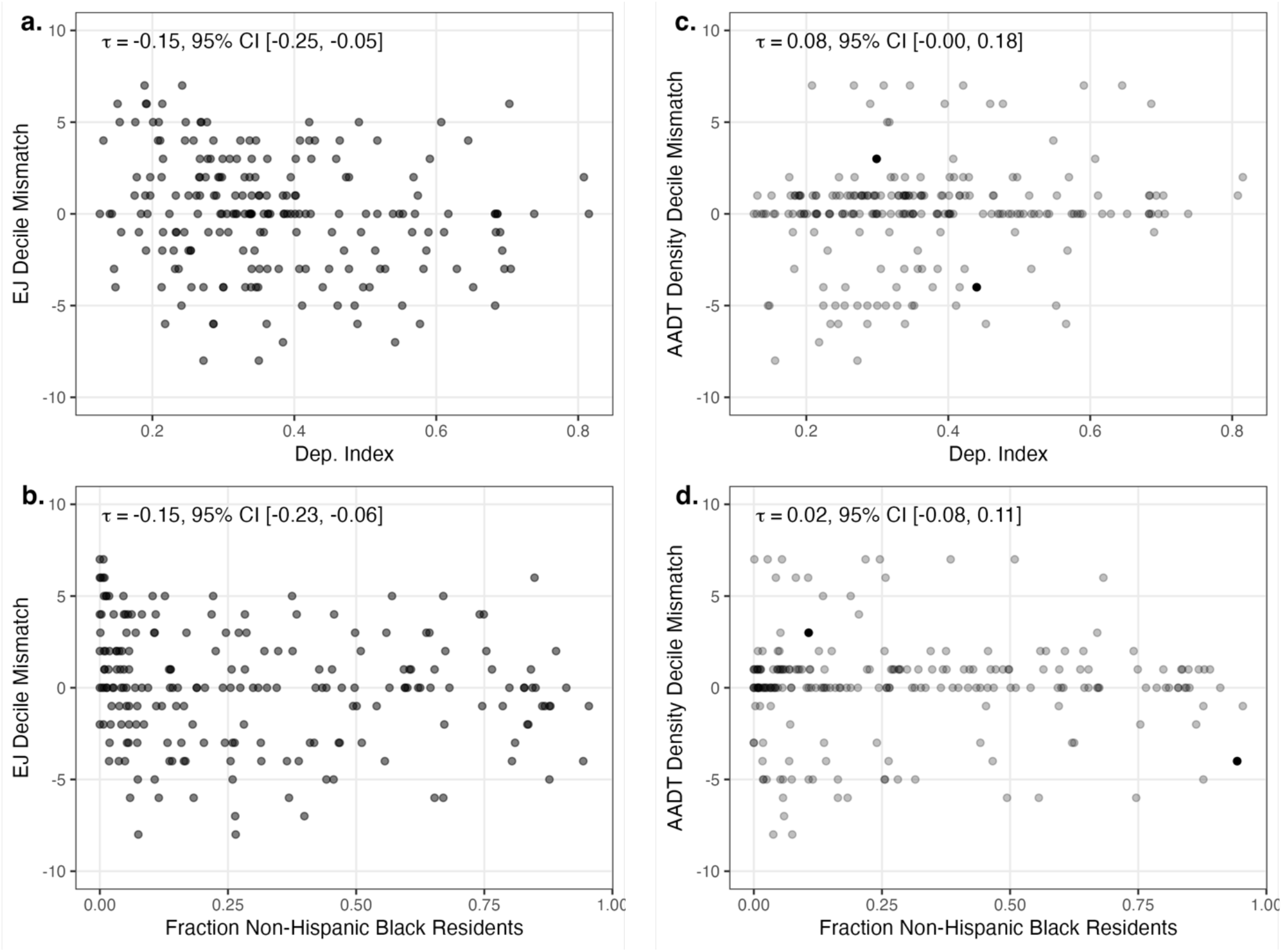
Scatterplots showing the relationship between the parcel-based bias of each census tract-level traffic measure with the material community deprivation index and the fraction of black residents, with tau(τ) representing the value of Kendall’s correlation coefficient.

## Discussion

While the errors were not biased against disadvantaged groups, the inconsistencies across sociodemographic measures can manifest in detrimental downstream effects as EJ Screen aggregation techniques may over-estimate exposure in predominantly black or materially deprived neighborhoods.

Clinically, this may lead practitioners to overtreat patients from highly deprived or predominantly black neighborhoods. Overtreatment can contribute to an unnecessary burden on the patients, and clinics themselves. This may be especially true for patients with respiratory conditions, such as asthma or cystic fibrosis^18^. This bias, when viewed through a research lens, may act as a suppressant of the known effects of pollution exposure. Through a research perspective, this bias might serve to downplay the recognized consequences of pollution exposure. Researchers could be led to believe that pollution has a lesser effect because of the assumed higher levels of exposure in underprivileged groups, leading to an oversight of a possible factor for adverse outcomes. Taken together, these consequences could result overtreatment from health systems, leading to negative psychological and sociological effects^19^, while concurrently suppressing potentially impactful research^11^. Lastly, it has been demonstrated that even correcting for potential misclassification provides diminishing returns, making it all the more important to accurately measure and classify exposures^20^.

In conclusion, we recommend investigators to develop their own parcel-based traffic pollution measure where available. In the cases where a parcel measure is not feasible, we recommend utilization of the AADT moving truck density at a relatively fine geographic level as it is likely to be fairer and more accurate than the EJ Screen traffic proximity. EJ Screen is often chosen by investigators because it is pre-computed, but we have shown that there are limitations to using the EJ Screen traffic proximity. Lastly, future research should consider downstream effects of unfair exposure assessments in clinical, policy, and research settings.

## Data Availability Statement

The data used in this analysis are publicly available in known repositories, as well as at the following web address: https://github.com/geomarker-io/parcel.

## Statements and Declarations

### Conflicts of Interest Statement

The authors have no conflicts of interest to disclose.

Funding provided by NIH grant 5R01LM013222-04

## Acknowledgements

Technical assistance, advice and support provided by all listed co-authors. Thanks to them for their support.

## Notes

### Competing Interest Statement

The authors have declared no competing interest.

